# The ‘Icarus effect’ of preventative health behaviors

**DOI:** 10.1101/2020.06.08.20126029

**Authors:** Simon Carrignon, R. Alexander Bentley, Matthew Silk, Nina H. Fefferman

## Abstract

Ongoing efforts to combat the global pandemic of COVID-19 via public health policy have revealed the critical importance of understanding how individuals understand and react to infection risks. We here present a model to explore how both individual observation and social learning are likely to shape behavioral, and therefore epidemiological, dynamics over time. Efforts to delay and reduce infections can compromise their own success, especially in populations with age-structure in both disease risk and social learning —two critical features of the current COVID-19 crisis. Our results concur with anecdotal observations of age-based differences in reactions to public health recommendations. We show how shifting reliance on types of learning affect the course of an outbreak, and could therefore factor into policy-based interventions.

## 2 Introduction

Among the research gaps in the response to COVID-19, declared a pandemic in March/Feb 2020 [1], a WHO strategic panel has called for “Comparative analysis of different quarantine strategies and contexts for their effectiveness and social acceptability” [2].

A familiar challenge is that the more successful a preventative strategy is in slowing an epidemic like COVID-19, the more public demand there is to relax those successful efforts before the epidemic has passed. This recalls the ‘Icarus paradox’, or failure brought about by the same strategy that led to initial success [3]. Successful preventative public health measures can facilitate the illusion that they were unnecessary in the first place.

For COVID-19, the challenge is intensified by its evident transmission in the absence of obvious symptoms [4]. Without adequate testing of a population [5], undocumented infections may outnumber documented infections by an order of magnitude [6, 4, 5]. This implies that (a) most people will underestimate the threat based on personal observations of infected individuals, and that (b) general social distancing in a population will be more effective than simply isolating or avoiding individuals with obvious symptoms. We expect to see these dynamics play out as groups continue to gather despite public health recommendations against it [7].

The behavioral dynamics can be complex [8]. Social distancing comes at an economic and psychological cost [9], yet the level of benefit (avoided risk) remains hidden, potentially until the epidemic has spread well beyond the point at which that cost would have been justified and/or acceptable. Encouragingly, social influence or conformity can lead people to adopt a costly beneficial behavior before individually observing direct evidence of its benefits [10, 11]. Subsequently, as the true benefits become more transparent to the public, individual cost-benefit decisions can support the behavior, in place of social conformity.

Understanding these dynamics are likely critical in designing successful on-going policies for ‘flattening the curve’ (Fig. 1), especially as local or national governments begin to reopen without extensive testing capacities. The aim is to tease apart the timing and impact of these different mechanisms for influencing individual behavioral choices over the course of an outbreak. If well-informed by likely dynamics in the factors that contribute to individual decision-making, policies may be able to forestall eventual rejection and remain more effective in the longer term.

**Figure 1:**
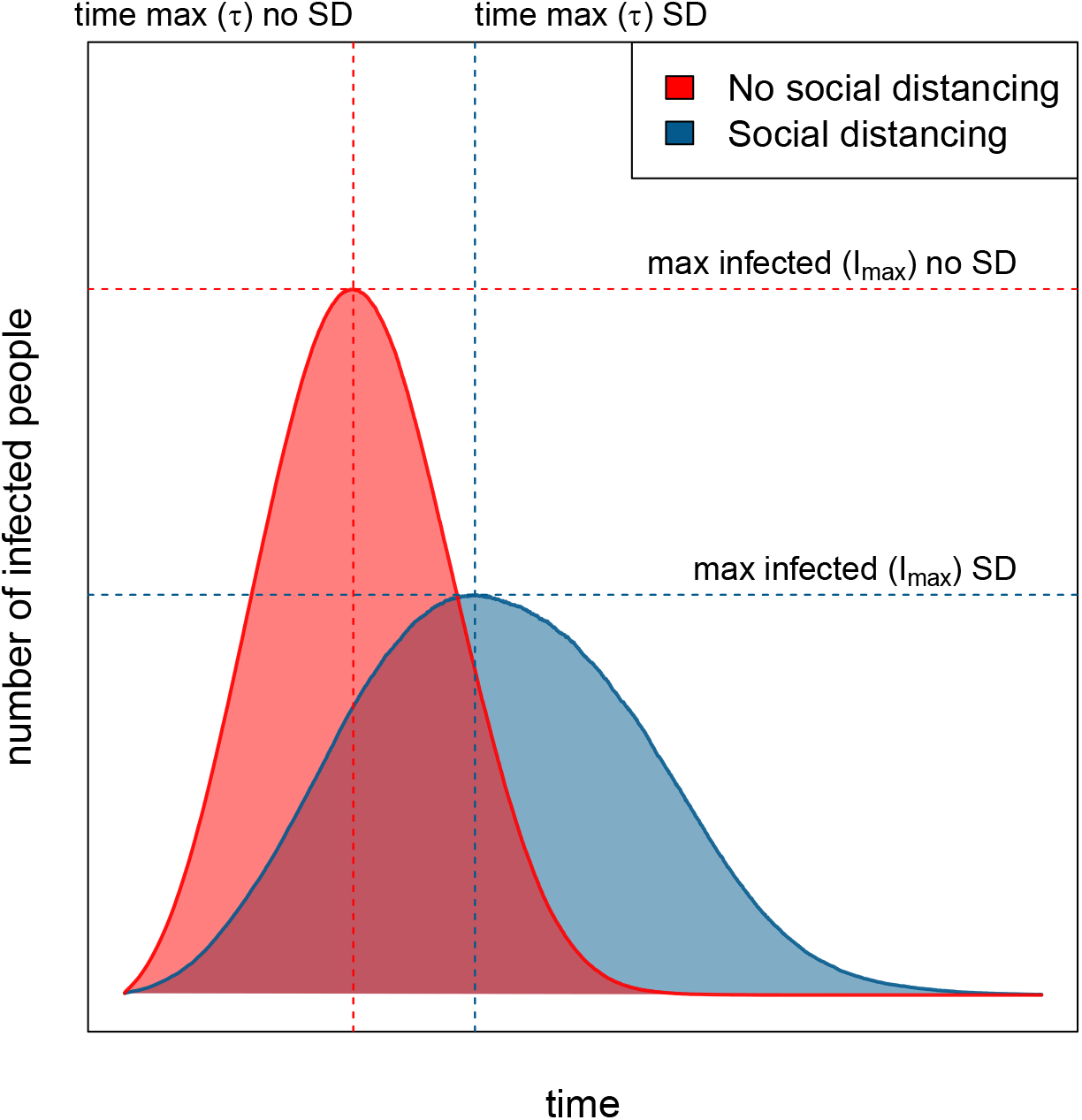
Illustration of the impact of social distancing on the spread of the virus.

Here we frame the impact of the ‘Icarus paradox’ as dependent on information and timing. At what point do individuals observe enough infections around them to adopt preventative behavior as their own cost/benefit decision? Before this point, in the absence of government directives, social norms may be needed to encourage protective behaviors as people are not yet observing many infections. We expect that early in an outbreak, the link between protective behaviors and being uninfected will not yet be transparent. As infection prevalence increases, and the benefits become more obvious to individuals, social learning becomes less necessary to facilitate preventative behaviors.

To explore these complex behavioral and disease dynamics, we employ an agent-based modelling approach.

### 2.1 Model rationale

Here we take the approach of discrete behavioral choice with social influence [12, 13, 14, 15], where we model decisions as based on a separable combination of two components: observational and social learning. Importantly, the underlying physical contact network through which a disease might spread [16, 17, 18, 19, 20], which can be mitigated by physical distancing, are different than the ‘contagion’ pathways of information, behaviors, or beliefs [21, 20]. Of course, information pathways include variety of media and daily communicative activities, whether physically close or distant.

#### 2.1.1 Observational Learning

The individual observational learning component is governed by the payoff difference between options, based on information people receive or observe to form beliefs about their health risks [22, 23, 10, 20]. For example, the perceived utility of social distancing likely increases as more illnesses and deaths are observed.

In typical formulations of discrete choice theory or quantal response theory, the probability that an agent chooses choice *i* at time *t* is proportional to *e*^*κU*^*i*^(*t*)^+ *∈*_*i*_(*t*), where *κ* is the transparency of choice and *U*_*i*_(*t*) is the intrinsic utility of the choice, and *E*_*i*_(*t*) is a noise term [24, 12, 13, 25]. Normalized across all choices, the shape of the probability function has a sigmoid shape with respect to the utility of the choice.

In our model, we therefore assume the probability of choice through observational learning takes on a sigmoid form, shaped by two parameters. One parameter for this is the point at which one option has a higher utility than the other, *U*_*i*_ *> U*_*j*_: we call this the ‘inflection point’, *ν*. In our model, this inflection point represents the level of viral infections people perceive (assumed proportional to the number of infections in the age cohort) that signals the positive utility of social distancing in reducing infection risk. This average inflection point is a convenient modeling assumption; in reality there would be a distribution among the population as well as an ‘identification problem’ in detailing how a group’s average behaviour influences its individuals [26, 27].

The second parameter for the observational learning component is transparency of choice, *κ*, which governs the steepness of the sigmoid curve: the higher the transparency of choice, the steeper the shape becomes, i.e. the smaller the variance in selecting the highest utility, *U*. In the real world, transparency might range from a clear, government-issued stay-home order at the fully transparent end of the spectrum, to a cacophony of conflicting media messages at the non-transparent end.

One way of effecting behavioral change is when an individual learns by themselves, by looking at the health status of people of the same age and estimating if they should switch to the other behavior. Given *I*_*a*_, the proportion of infected individuals in the same age cohort, *a*, the probability for an individual *i* to switch from ‘non-adherent’ status, *NA*, to ‘adherent’ status, *NA*, is defined as:

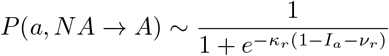

where *κ* represents the transparency of choice—determining the steepness of the sigmoid curves—and *ν* defines the point at which the individual has a 50% probability to switch to adherent status, *A*. The influence of *ν* and *κ*, respectively, on the probability of adoption, *P*, is illustrated in Figure 2.

**Figure 2:**
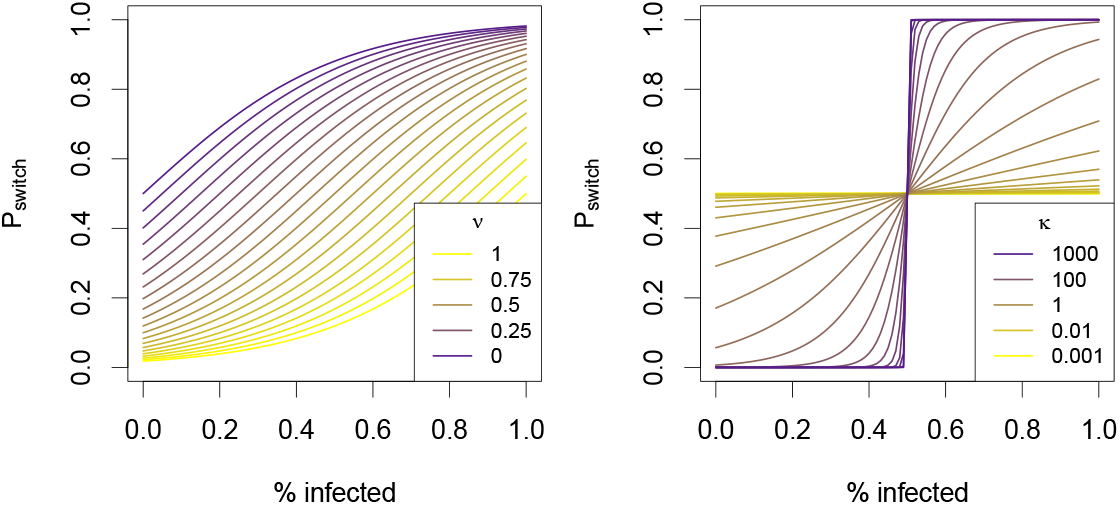
Illustration of the impact of transparency of choice, *κ*, and point of inflection, *ν*, on the decision curve.

The same function is used to calculate the probability for an individual to switch back from adherent to non-adherent behavior given the fraction of people in the age cohort who are not infected, (1 − *I*_*a*_):

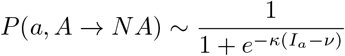

where *ν*_*r*_ is inflection point for reverting to *NA* behavior (and can be different from *ν*).

#### 2.1.2 Social learning

Social interactions play an important “role in mediating the spread of social contagions that impact health outcomes,” [20]. To simplify their complex role for our model, we employ the useful distinction between complex beliefs versus ‘simple’ social contagions [20]. Here we treat social distancing and related protective behaviors as simple contagions: copying what others do. In our model, protective behaviors can be learned easily and quickly from others. For example, people may wear a mask to the supermarket because they saw other shoppers wearing masks. For our scale of modeling aggregation, we assume social influence operates ‘as if’ each person copies a currently healthy person, randomly encountered in the population, within a certain social or physical distance. The effect is the frequency of copied behaviors is stochastic, where probability of selecting option *i* is proportional to its current frequency or popularity, *p*_*i,t*_ [12, 13, 26].

We also assume that social learning of simple behaviors (as opposed to complex skills learned over years) is age-restricted, in that people will only copy others in their own age cohort. For convenience, this assumption conflates two observations, that: (a) age cohorts tend already to share similar beliefs and preferences, derived from both ontogeny and broadly shared socio-economic landscape during early years of development [28, 29, 30, 31, 32, 33, 34], and (b) conformity tends to be age-dependent with age-biased social learning [35].

### 2.2 Agent Based Implementation

We now present an agent-based model (ABM) that explores two related dynamics. Good protective behaviors reduce the transmission of the disease, whereas observing other infected individuals encourages (through observational learning) the adoption of protective behaviors.

The ABM consists of agents who are involved in two inter-related processes, the contagion of a disease in SIR fashion [36], and the spread of protective behaviors through a combination of individual and social learning. Agents move in a grid, *x* units by *y* units in dimension, with a normally-distributed range of speeds. An infected agent can infect another agent if they are within distance *d* of each other. At the the same time, agents of similar age are observing the levels of infection as well as protective behaviors, and adjusting their behaviors according to the observational and social learning rules described above.

The algorithm describing the details of the model execution an be found in the supplementary material. There are two main functions, one (*generatePopulation*) generates the initial population of agents and the other (*selectModel*) allows agents to copy the behavior of another agent, who must be both non-infected and within the same age cohort, within radius *r* of the agent.

Simulations were run with the parameters described in the Table 1 with the population initialised as described by the algorithm (see supplementary materials). Some parameters were fixed (*N, d*_*i*_, *i*_0_), behaviors (*B*_0_) were all set to NA initially, speeds were sampled from a normal distribution *S* ∼*N* (1, 0.2) and recovery times were sampled from a uniform distribution *R*∼ *U* (8, 14) × 24. The parameters were selected such that, overall, agents encounter a mean of 12 individuals per 24 time steps, such that each time step represents one hour. The recovery times in the SIR model were thus calculated to represent 8 to 14 days as a reasonable approximation for COVID-19 [37].

**Table 1:** Parameters used in the agent-based simulation. The Algorithms for the simulation are in the Supplementary material.

| parameter | description | value |
| --- | --- | --- |
| $N$ | number of agents | 500 |
| $P(b)$ | probability to transmit disease w/ behavior $b$ | $[p_{NA} = 1, p_A = .1]$ |
| $p$ | proba. to learn by individual observation | $U(0, 1)$ |
| $r$ | radius of social learning | $U(1, 50)$ |
| $t_{step}$ | number of time steps | 1500 |
| $d$ | maximum distance for disease transmission | 5 |
| $i_0$ | number of initial infection | 1 |
| $\nu$ & $\nu_r$ | inflexion point of the learning functions | $U(0, 1)$ |
| $\kappa$ & $\kappa_r$ | transparency of choice for adopting & reverting | $10^{U(-1, 3)}$ |
| $x$ & $y$ | size limits of the space | 100 |
| $R$ | distribution of recovery times | $R \sim U(8, 14) \times 24$ |
| $B_0$ | initial distribution of behaviors | $B_0^i = \text{NA}, \forall \text{ agent } i$ |
| $S$ | distribution of the speeds of the agents | $S = N(1, 0.2)$ |

To explore the impact of learning on the disease outcomes over the course of an outbreak, we ran three sets of models. We first ran two baseline conditions in which individual behavior is held constant as either non-adherent (worst case scenario) or adherent (best case scenario) over the course of the simulation. We subsequently ran the models with the parameters described in Table 1 after initializing the population following Algorithm2. In all, we ran 826 892 simulations and recorded the number of infected agents at each time step in each simulation. To summarize our simulations in a way that captures the ‘flattening the curve’, we focused on two metrics: (a) the maximum number of infected people *I*_*max*_, and (b) the time to reach this maximum *τ*. As in Figure 1, the ‘flat’ curve has lower *I*_*max*_ and larger *τ*. Across all simulations, we recorded the largest and smallest maximum total infections among all the runs, as max(*I*_*max*_) and min(*I*_*max*_), respectively, as well as the longest and shortest times to reach the max, as max(*τ*) and min(*τ*), respectively. Since these dimensions will tend to be inversely correlated, we defined a metric, *δ*(*s*), for a set of simulations *s* summarizing both:

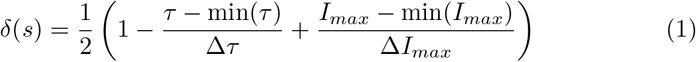

where Δ*I*_*max*_ = [max(*I*_*max*_) −min(*I*_*max*_)] and Δ*τ* = [max(*τ*) −min(*τ*)], from all simulations.

Whereas our results below involve the simulation just described, we also explored three alternative scenarios, described in the Supplementary Information. In the first scenario individuals could copy any other individual via social learning, rather only those seen to be healthy. In the second scenario, part or all of the initial population already adhere to social distancing. The third scenario allows learning only after a certain number of time steps (see Supplementary Information).

## 3 Results

Our simulations produced meaningfully different outcomes that varied in the success of ‘flattening the curve’ (Figure 1). This reflects the expected patterns of outbreak curves under the impact of social distancing.

Which learning parameters yielded the ‘flattest’ curve? The simulations reveal a mix of parameters underlying the best preventative outcomes. Among the simulated outbreaks in which *δ* were minimized—what we call the ‘best’ outcome—the transparency and inflection points (*ν*_*r*_ and *κ*_*r*_) for reversion to non-protective behavior needed to be more constrained than the respective parameters (*ν* and *κ*) for choosing protective behavior. Figure 5 shows that the range of optimal parameters are more constrained by the end of the simulation (bottom row), which represents about two months, than they were after just 150 time steps (top row), representing approximately one week (see gray line in Figure 3). Supplementary Figure 9 shows these joint posterior distributions in more detail.

**Figure 3:**
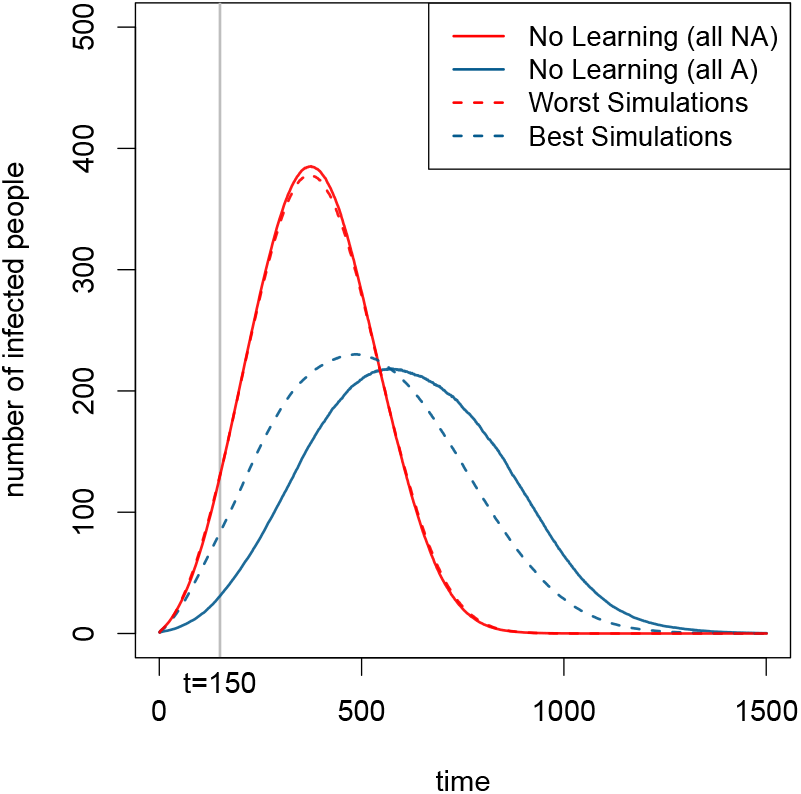
Comparing the mean trajectories of SIR model where agents do not learn and instead continue in their initial behavior, with trajectories generated by simulations. Red curve: all agents are non-adherent to social distancing and stick to this behavior. Blue curve: all agents adhere to social distancing. Dashed red line: mean trajectory of the 1, 000 worst simulations. Blue dashed line: mean trajectory of simulations run with the parameters selected in the 1, 000 best simulations with regard to *δ*. Vertical gray line: status after 150 time steps.

**Figure 4:**
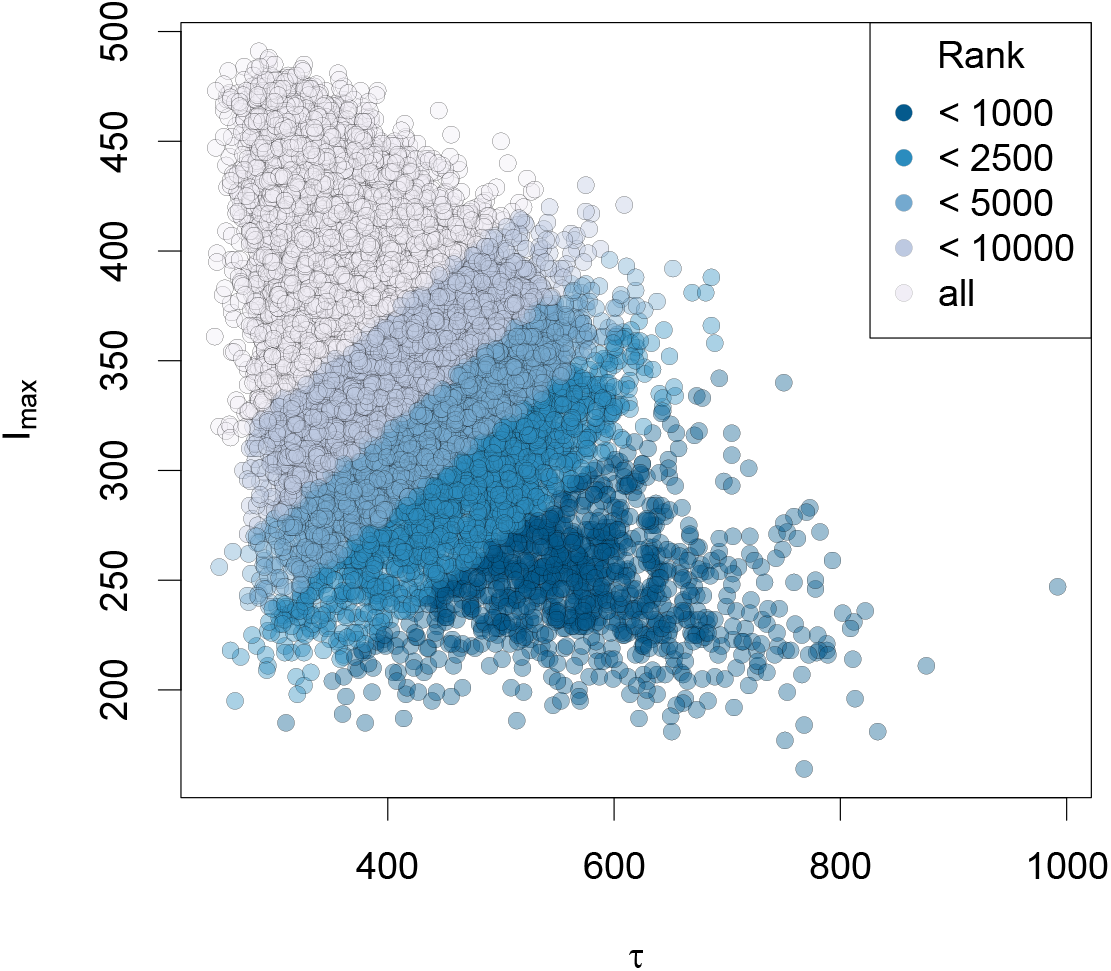
Distribution of 826, 892 simulations given two different metrics: the maximum number of infected people (y-axis) and the time to reach this maximum (x-axis). The colors represent different class of the simulations ranked given *δ*, the metric defined in Equation 1.

**Figure 5:**
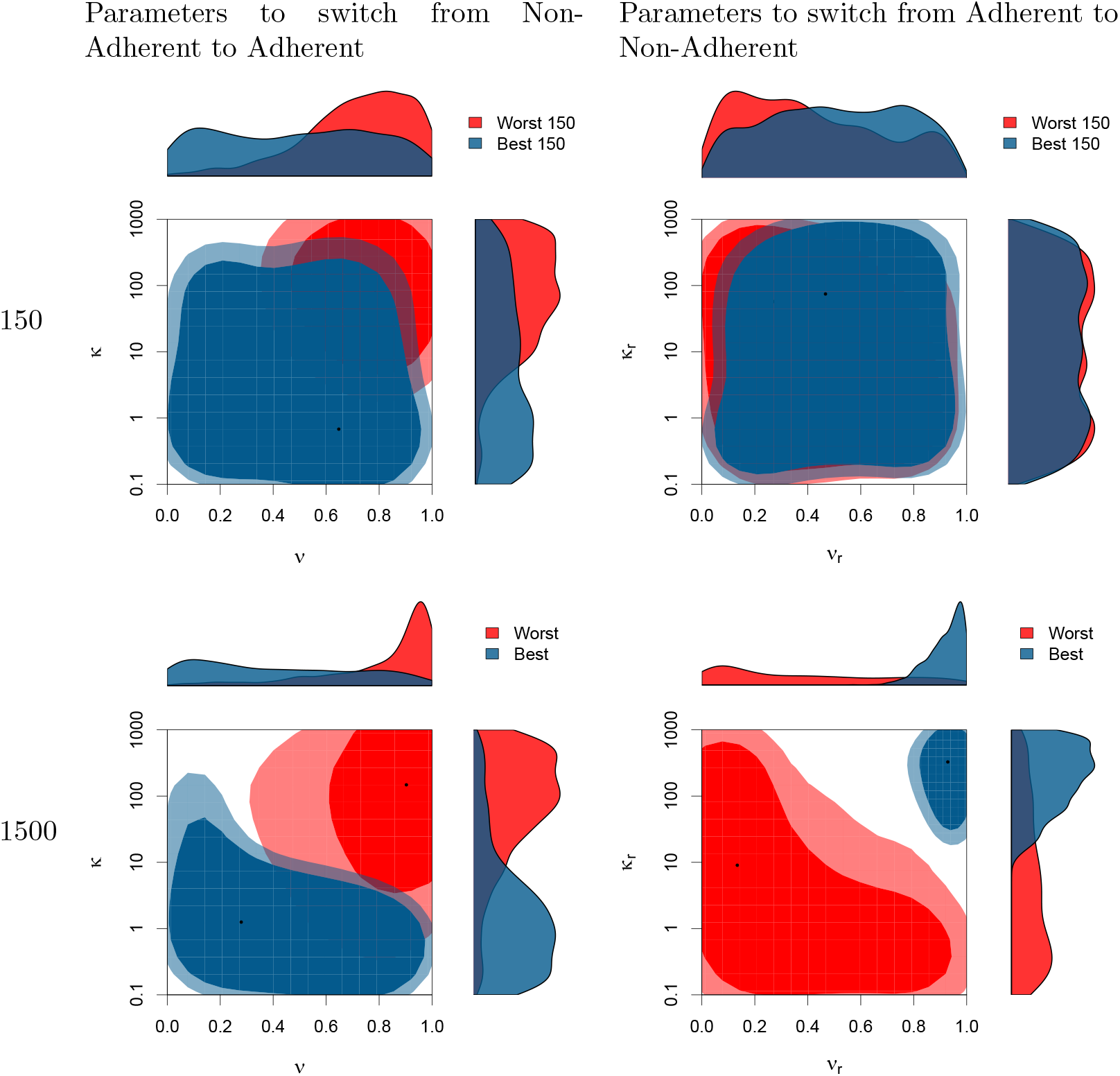
Joint posterior distributions for the parameters used to switch from Non-Adherent to Adherent (left column) and for the parameters used to switch back from Adherent to Non-Adherent (right column). The 2d areas represent the 70% and 90% HDR (High Density Region) *ie*, the smallest areas within which respectively 70% and 90% of the parameters combination fall (for the mathematical definition of HDR and how they can be represented see [38]. Lighter colors represent the 90% HDR whereas darker represent the 70% HDR. The top row represent the value for those parameters that minimize (in blue) or maximize (in red) the number of infected people at time step 150. The bottom row shows the parameters that minimize (in blue) or maximize (in red) *δ* (as defined in Equation 1) at the end of the simulation (1500 time steps). The marginal posteriors for each parameter taken separately are drawn in the margin.

Focusing on the end of the simulation (Figure 5, bottom row), we see different ranges of parameters yielding the ‘best’ (blue) and ‘worst’ (red) outcomes. Figure 5 shows with the joint posterior distributions of *ν* versus *κ* (adopting protective behaviors) and of *ν*_*r*_ versus *κ*_*r*_ (reversion back) on the left and right, respectively. Note particularly that the parameter regions flip-flop between adopting and reverting back. In other words, for the ‘best’ outcome, it is better to have low *κ* for adopting protective behaviors but then high *κ* for reverting back to non-protective behavior (see the cross-sectional distributions along the axes of each bivariate plot in Figure 5, lower row). At the same time, for the ‘best’ outcome *ν* can be virtually any value for switching to protective behaviors, but needs to be quite high for reverting back.

As a qualitative interpretation of the asymmetric effects of *ν*, the desired ‘flat’ curve is achieved through *low* transparency of choice in adopting the protective behavior, but high transparency about whether to revert back. Qualitatively the same is true of the inflection point, *κ*, which needs to be low in adopting the behavior but high in reverting back, in order to yield a ‘flat’ infection curve.

These results suggest the ‘best’ outcomes involved a ‘ratchet’ effect – a low, fuzzy barrier to adopt the behavior but a high, sharp (transparent) barrier to revert back. The low barriers to entry is consistent with theory on diffusion of innovations [39, 40, 41]

## 4 Discussion

Our results imply that combined observational and social learning can drive successful mitigation strategies (Figure 3). In an ‘Icarus’ scenario, where an initially successful strategy can lead to its own failure, the timing of these forms of learning are crucial. Our simulations reveal multiple possible outcomes of behavioral mitigation, from ‘flat’ infection curves to full, unchecked outbreak (Figure 4).

Those simulations yielding a flat curve generally required either low transparency of choice, low inflection point for adopting the behavior, and/or high inflection point for reversion back to non-protective behavior—in other words, readily adopting protective behavior and reluctance to give it up (Figure 5). It is not surprising that such risk-averse behaviors would protect a population. The challenge is that the risk needs to observable. Hence protective behaviors may be not be adopted until infection rates are very high, especially if most symptomatically infected individuals are not be publicly visible (e.g., remaining at home or in hospital).

For this reason, low transparency of choice—such as poor or conflicting information —can ‘jump-start’ adoption of protective behaviors by stretching out the inflection into a range, such that some individuals ‘mis-estimate’ infection risks as enough to trigger their decision (see Fig. 5). However, this result relies on the breadth of the distribution of responses to low transparency, rather than an alternative case in which either leadership or social norms cause low transparency to lead to greater average hesitancy to take any action [42].

Social learning dynamics were also crucial to outcomes. In social learning theory, “copying recent success” is often the best strategy [43]. If the disease symptoms/prevalence are transparent, copying healthy individuals (‘success’) should increase protective behaviors as disease prevalence increases. Because COVID-19 can be asymptomatic, however, transparency in this respect may be low. Infected individuals may seem healthy, such that non-protective behaviors can be copied even through a “copy success” strategy. This lack of transparency may critically compromise our ability to rely on learning-only strategies for successful disease risk containment.

These effects may be heightened among young people, for whom COVID-19 infection is more frequently asymptomatic and who are also socially influenced by peers of similar age [35]. Hence, while lower fatality risk from COVID-19 may help explain why some groups of young people rejected social distancing early in the U.S. outbreak [7], another important dynamic was likely social conformity and lack of transparency about infections among their peers.

One limitation in our simulations is that we employed a broad definition of observational learning, as agents estimate infection rates with some degree of precision (transparency). Observational learning is heterogeneous; at any given moment, each person observes different information and a different segment of the population. The real world also encompasses myriad information sources, individual experiences, and biases. Also, information from media will have a different effect on decision-making than having friends and family become ill [44]. Individuals who follow stay-home protocols have different interactions than those who interact in public spaces. Taken together, these processes affect the transparency of how observably protective behaviors relate to disease risks.

These considerations are particularly relevant for mitigating pandemic spread where strong governmental control is not possible. When individual choice is the driving factor in protective behaviors, age cohort effects become important since different demographic segments of societies are likely to be more initially risk-averse than others. In particular, older individuals may be early adopters of adherent behaviors because news reports of mortality rates in older populations create a psychological burden of fear. By contrast, younger and/or more economically limited individuals may delay switching to behavioral adherence, doing so only when they feel their circumstances allow it. In such cases, the socioeconomic and demographic representation of a population may be the critical drivers in determining whether individual behaviors, guided by learning, can be relied upon to effectively achieve outbreak mitigation by adherence to social distancing.

In our models, we have also assumed that choices are rational and focused only on epidemiological risk. In reality, social distancing involves considerable social and economic costs [45]). Human decision-making is also never fully rational, especially during period of stress [46]. Further, we have here explored only one potential route of social learning, in which individuals simply copy the behaviors of perceived healthy individuals. More nuanced approaches may emphasize preferentially copying people who share your beliefs, alignment with deeply-held beliefs or values, and other social learning strategies [47, 48, 49, 20]. Similarly, we have construed frequency of contact within a spatial radius to be the only medium for social derivation of learning. In reality, of course, individuals have many means of arriving at their sense of what other people are doing, including unique personal experiences and personal choice of media, however complicated or accurate [20, 50, 51, 52, 20]. This raises many interesting questions, such as the different effect of centralized versus diverse media, for example. Given our parsimonious starting point, future expansion and testing of our models can address these potentially important features of social groups.

## Conclusions

Understanding how the timing and dynamics of different types of learning affect individual behavior over the course of an outbreak, as disease prevalence changes the transparency of benefits of protective behaviors, while those behaviors become more visible as they proliferate. Social learning can help boost protective behaviors, but not until the number of infections has brought about those behaviors initially through observation-driven decision making.

These considerations may be critical in shaping policies that will foster public adherence. True leadership must sometimes accept the burden of enacting policies doomed to be unpopular. However, understanding of the role of the ‘Icarus paradox’ in public health safety may help preemptive design of policies that anticipate increasing non-adherence as they are increasingly effective.

## Data Availability

All data will be made available upon request – as this is a modeling study, all data is the result of realizations of the computation described in the text.

## Acknowledgements

This material is based upon work supported by the NSF under award #2028710.

